# Visualizing the COVID-19 pandemic in Bangladesh using coxcombs: A tribute to Florence Nightingale

**DOI:** 10.1101/2020.05.23.20110866

**Authors:** Hasinur Rahaman Khan, Tamanna Howlader

## Abstract

Following detection of the first confirmed case of COVID-19 in early December 2019 in Wuhan, China, nearly six months have passed and almost every country in the world is battling against the COVID-19 war. The frontline warriors, namely the doctors, nurses and healthcare staff, have in many countries struggled to care for the sick under conditions of limited resources and protection and the threat of an overwhelmed healthcare system. It is during times such as this, that we draw strength and inspiration from Florence Nightingale - a passionate statistician, social reformer, feminist champion and a pioneer of modern nursing and data visualization. Nightingale’s famed Florence Night-ingle Diagram also known as “coxcomb”, which was created 150 years ago and used to display the causes of death in the British Army hospital barracks, demonstrated how data visualization techniques could be a powerful medium of communication and a force for change. This paper pays tribute to Nightingale’s work by using data from Bangladesh to show that the coxcomb graph is still relevant in the era of COVID-19. The coxcomb graphs that have been produced to display COVID-19 data have provided deeper insights into the trends and relative changes of variables over the course of the pandemic. The paper also describes codes that allow one to easily reproduce the graphs using the statistical programming language R.

## 1 Introduction

In today’s data centric world, researchers from all disciplines have come to recognize the value of data visualization in its ability to tell the story in the data. Data visualization has become an indispensable tool in any scientific investigation, report and presentation because it can effectively communicate information in data in a visually appealing an easy-to-understand manner. In the era of Covid-19, data visualization is particulary relevant because it is helping to shape public understanding of the pandemic. It is being used to both inform the public and policy makers as well as to persuade people to change their behaviours. The literature mentions several individuals such as Edward R. Tufte, William S. Cleveland, and Cole Nussbaumer Knaflic [(Bounthavong, 2020), (Fee & Garofalo, 2010)], who have made significant contributions towards advancing data visualization. But among them all, Florence Nightingale deserves special mention. Florence Nightingale (1820-1910) was a nurse, statistician, and social reformer. Mathematicians and data scientists revere Nightingale as one of history’s most important statisticians and a pioneer of data visualization. Not only did she use data for forcasts and comparisons to find the causes of problems (Hoyos, Accessed May 15, 2020), but she also demonstrated the power of data visualization as a force for change at a time when war and public health became important partners in improving healthcare.

During the Crimean War, which broke out between Britain, France, Sardinia, Russia, and the Ottoman Empire during 1853-1856, wounded British troops were shipped across the Black Sea to hospitals in Turkey. More than 21,000 British troops died but only 5,000 deaths were attributable to actual battle. Nightingale was relentless in her pursuits to determine why troops were dying in these hospitals. When she was appointed Superintendent of the Female Nurses in the British Military Hospital Barracks in 1854 by Sydney Herbert, the then Secretary of War, she brought with her a team of 38 volunteer nurses and an innovative and determined mind (Fee & Garofalo, 2010). Nightingale began implementing reforms in the hospital barracks. She instituted sterilized laundry and hand washing sanitation protocols, raised funds, and improved hospital administration. Moreover, during her tour in the Crimean War, Nightingale collected an impressive collection of data about mortality in the army. When she returned to England after the war, Nightingale befriended William Farr, a physician, statistician, and data visualization pioneer. He helped her recognize the potential of the data she had assembled in Crimea. Nightingale went about describing the data in visual detail as an appliied statistician would.

In 1958, Nightingale submitted a report to the Royal Commission on the Health of the Army from her statistical records of the casualties during the Crimean War (Archive, Accessed May 15, 2020). The report was illustrated with a new type of pie diagram called the Nightingale rose or wedge diagram (Figure 1), which later came to be known as the Florence Nightingale diagram, coxcomb and polar area diagrams (Understanding Uncertainty, Accessed May 15, 2020). The Florence Nightingale diagrams that appeared in the report were constructed using Table (1), which was also published in the same report (Archive, Accessed May 15, 2020). The diagram was made up of several segments or petals where each petal represented a month. The diagram took advantage of the radii of the segments or petals in addition to their length from the center to generate areas that reflected the scale and size of the estimated mortality rate (deaths per 1000 population) of soldiers in different months. It also enabled one to discern the scale of the mortality by month relative to other months based on the area. From these graphs it was immediately evident that most of the soldiers were dying not because of combat, but due to common camp diseases, which were mitigable. This type of visual aid prompted the military to review how the soldiers were being treated and led to reform that helped to reduce noncombat related mortality in the British Army. In 1860, Florence Nightingale created the Nightingale Training School at St Thomas Hospital (now called the Florence Nightingale Faculty of Nursing and Midwifery and Palliative Care at Kings College London) to train a new generation of nurses using her ideas and philosophies. Her accomplishments in nursing, public health, and statistically-driven ideas of social reform made her a hero and in one of his poems, Henry Wadsworth Longfellow immortalized her as”The Lady with the Lamp”.

**Figure 1:**
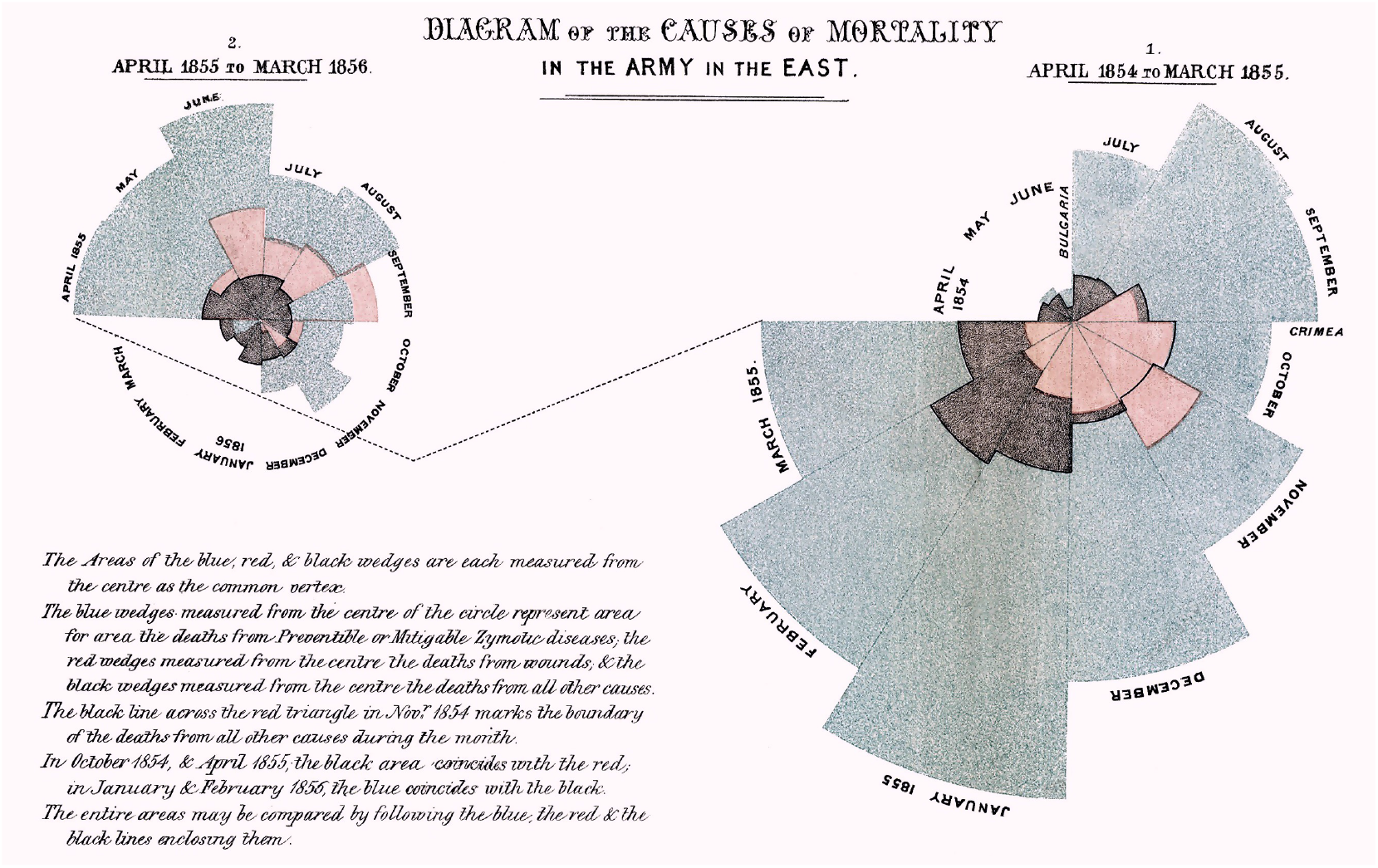
Florence Nightingale rose diagram illustrated the causes of death in the British Army. 1858.

**Table 1:**
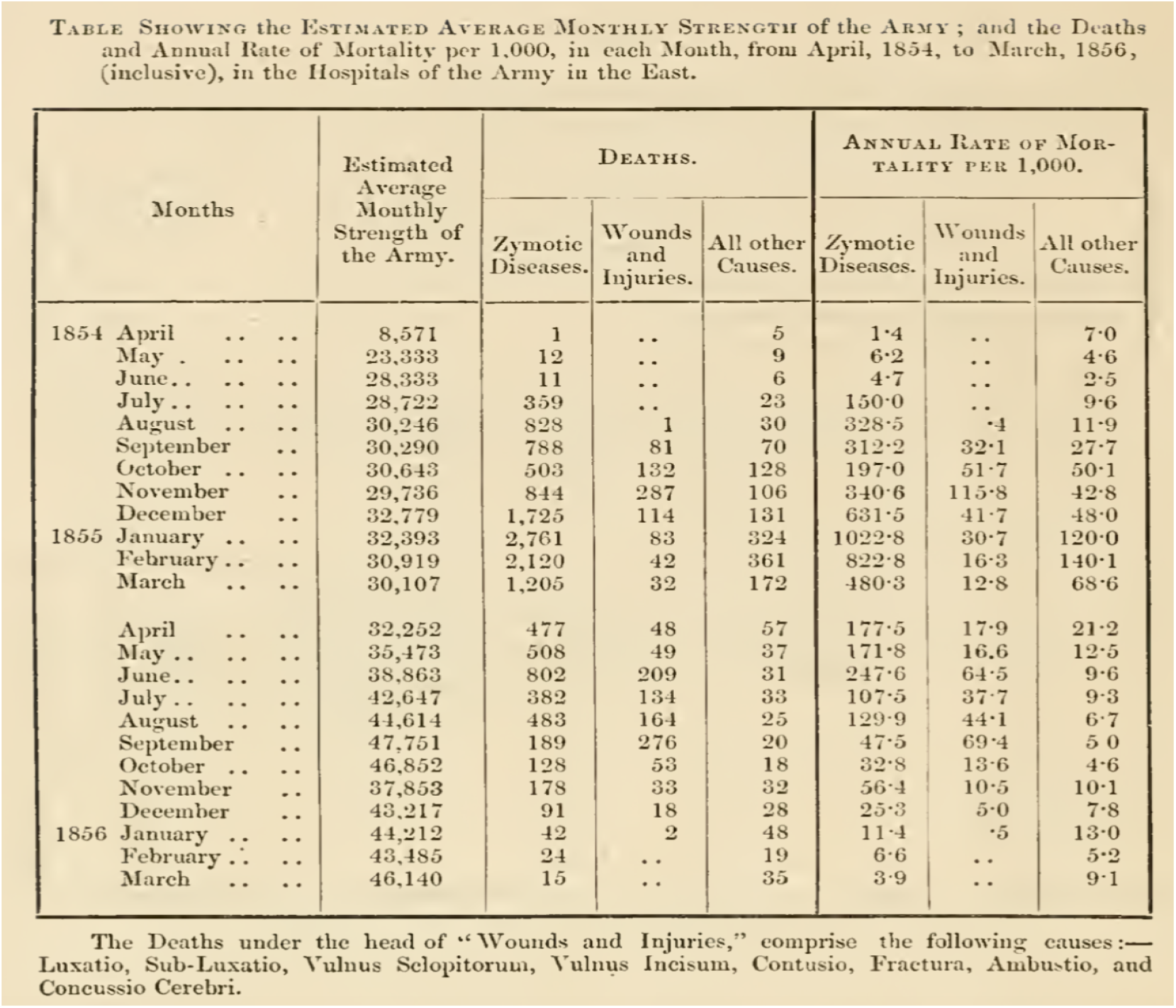
Estimated Average Monthly Strength of the Army; and the deaths and Annual Rate of Mortality per 1000 in each month, from April 1854, to March 1856.

Florence Nightingale’s story will be told by generations. In as early as the Victorian age, Nightingale demonstrated how powerful data visualization can be in influencing change when intelligently used. Apart from the famous coxcomb diagram, she also created bar charts, stacked bars, honeycomb density plots, and 100% area plots (Understanding Uncertainty, Accessed May 15, 2020). She was obsessed with data quality and standardization. She orchestrated multi-year data collection efforts across the world using model survey forms of her own design. Florence Nightingale has changed the way we use and view data and her work continues to inspire statisticans even till this day. She was the first female member of the Royal Statistical Society and an honorary member of the American Statistical Association [(Nightingale, Accessed May 15, 2020), (Andrews, Accessed May 15, 2020)]. More information on Nightingale is available in Andrews (Accessed May 15, 2020), Hoyos (Accessed May 15, 2020), Johnson (2008), The Economist (Accessed May 15, 2020), Small (1998), Lienhard (Accessed May 15, 2020), Blog (Accessed May 15, 2020), Rogers (Accessed May 15, 2020), HistoryInformaation.com (Accessed May 15, 2020), Datylon blog (Accessed May 15, 2020), BBC (Accessed May 15, 2020).

In modern times, Florence Nightingale’s coxcomb diagrams are still being used to visualize data in different applications. For example, Lockwood (Accessed May 15, 2020) demonstrated the effective use of coxcomb diagrams with sports data. More specifically, the authors generated various coxcomb graphs for 2017 English premier league statistics. Only recently, have some researchers begun to see the value of the coxcomb diagram in depicting temporal trends of COVID-19 data. For instance, coxcomb diagrams have been constructed for number of daily confirmed cases, the proportions of active cases, number of recovered patients and number of deceased across a map in (Field, Kenneth, Accessed May 15, 2020). The authors emphasized the benefits of the technique in representjng an immense amount of information in an efficient and elegant design. The objective of the current study is to construct coxcomb diagrams to understand the COVID-19 pandemic in Bangladesh. In particular, it examines the temporal patterns in the daily rates of infections, deaths and recovery for Bangladesh over a span of 10 weeks using coxcomb diagrams. In addition, this article describes in detail the R-codes needed to generate coxcomb diagrams in the hope that it will inspire others to glean valuable information from their data using this excellent data visualization tool.

## 2 Methodology

The data on COVID-19 in Bangladessh has been collected from worldometers.info website (Max Roser & Ortiz-Ospina, 2020). We considered data reported as of May 16, 2020. The data has been cross checked with the Bangladesh government’s source (Institute of Epidemiology and Disease Control and Research, 2020). The severe acute respiratory syndrome coronavirus (SARS-CoV-2) pandemic fully established itself in Bangladesh in early April when the number of infected persons crossed the 100-th mark. Prior to this time, the country detected its first three coronavirus cases on 8 March 2020 (Institute of Epidemiology and Disease Control and Research, 2020), (Khan & Hossain, 2020b), (Khan & Hossain, 2020a), (Khan et al., 2020). Since early April, the number of new infections continued to rise as Bangladesh ramped up the number of tests performed. By 16th of May, the country counted 10229 cases and a death toll of 183 persons (Institute of Epidemiology and Disease Control and Research, 2020), (Khan, 2020).

We explain here how the original coxcomb diagram shown in Figure 1 (Hoyos, Accessed May 15, 2020) was constructed by Nightingale. The segments in these coxcombs were the months of the year. There are twelve months in the year, so the angle of each sector is, in degrees,

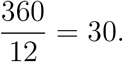

The data represented by the coxcombs are deaths each month for British soldiers in the Crimean War. We reproduce here the mathematics that Nightingale used in order to calculate the data for deaths by diseases (the blue portion of the Figure 1). Suppose that on average during a particular month there were *S* soldiers, and *D* of these soldiers died of diseases during that month. Nightingale considered the proportion of soldiers who died of diseases; that is, she considered the quantity 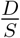. Next, she wished to express this quantity as a figure out of 1000, so she multiplied it by 1000, to obtain 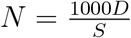. Thus, in that month, N out of every 1000 soldiers died of diseases. Finally, Nightingale multiplied N by 12 to indicate how many soldiers would have died from diseases throughout the whole year had the circumstances of that month been replicated throughout the year. She described the final quantity that she obtained as the annual rate of mortality per 1000 and she calculated this quanity for each month of the year. It is given by the formula

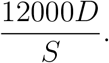

This is the value of the area of the blue sector for the particular month that we are considering. Of course, area depends on the unit of measurement, so Nightingale will have scaled all her values appropriately. Since coxcombs do not have scales, Nightingale need not have multiplied 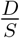 by 12000 in order to plot the coxcombs.

## 3 Analysis and Results

### 3.1 Analysis of COVID-19 Data Using Coxcombs

The COVID-19 dataset for Bangladesh contains nine variables, which are defined as follows:‘Date’ represents dates from date of first case detected (March 8, 2020) to May 16 under the format ‘year-month-day’; ‘Week’ represents the weeks coded as 1-10 weeks; ‘Month’ represents the months coded as March, April and May; ‘Daily deaths’ represents the number of daily reported deaths;‘Daily recovered’ represents the number of daily recovered individuals from COVID-19; ‘Daily cases’ represents the number of daily reported infections;‘Daily deaths.rate’ represents the rate of daily reported deaths per crore; ‘Daily recovered.rate’ represents the rate of daily recovered individuals from COVID-19 per crore and ‘Daily cases.rate’ represents the rate of daily reported infections per crore. It may be mentioned that the rates per crore were obtained by dividing the daily data by 17 since the total population in Bangladesh is 17 crore or 170 milion.

We are interested to produce coxcomb graphs for the rates variables. Before constructing the coxcomb graphs, we obtained first the stacked barplot, as shown in Figure 2, for the three rate variables. The graph is difficult to read when the number of days increases so that for a long time series data, the stacked barplot is not very useful. Furthermore, the daily death rates are not visible due to their low values. Although this graph gives a clear picture of the trend for rate variables, it is quite difficullt to compare the week-wise rates. We created a coxcomb diagram against this stacked barplot (see Figure 3). It is immediately evident that the rate of daily reported infections per crore picked up rapidly after the 4th week since March 8, 2020, and continued to increase in successive weeks.

**Figure 2:**
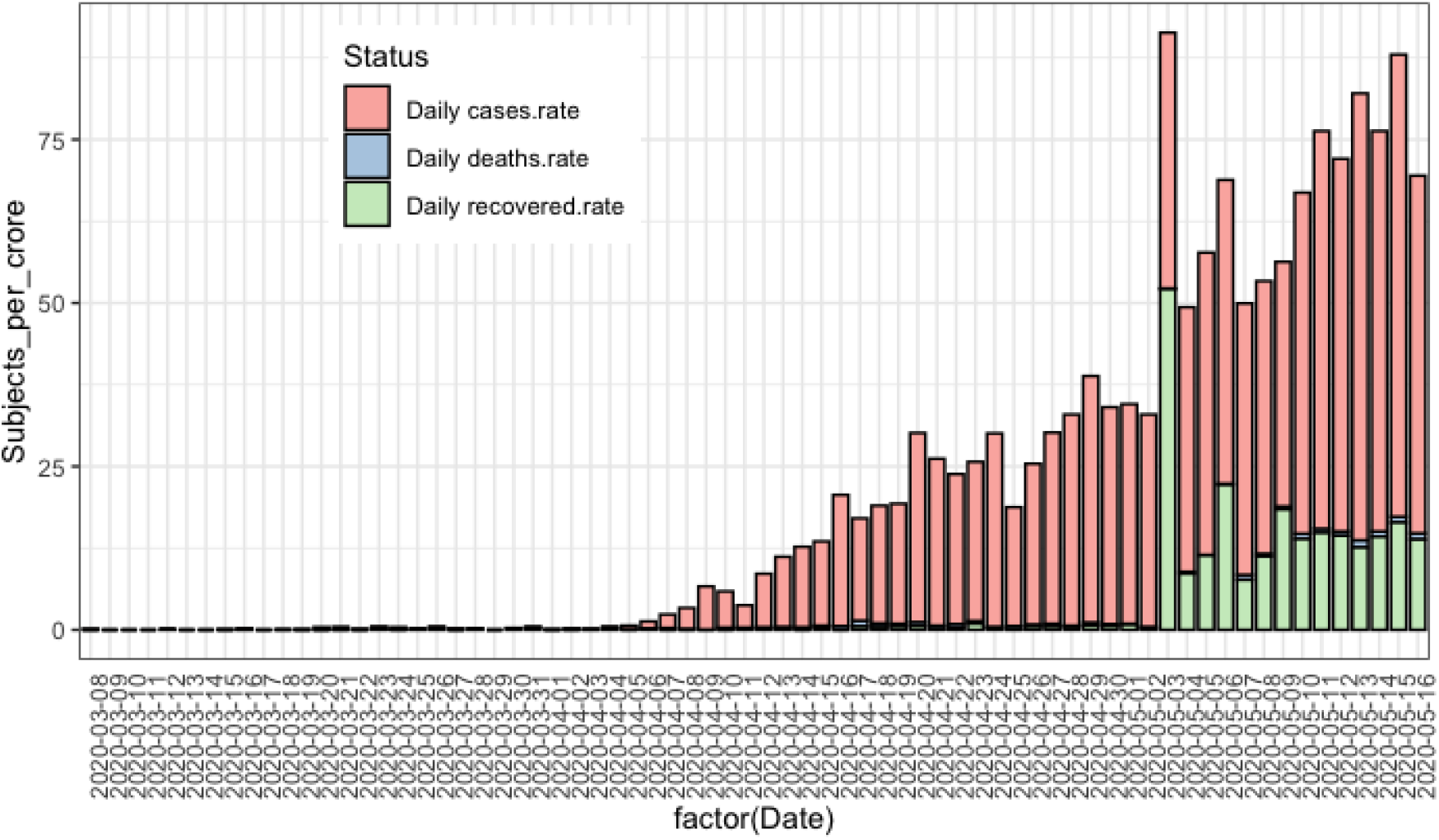
Stacked barplot for daily rate of infections, deaths and recovery for Bangladesh.

Thus, the coxcomb diagram is quite useful to compare the rates between the weeks and to trace the trend of the numbers. However, the statistics are not visible for the first four weeks because of their low values. It can be mentioned here that in first four weeks, the total number of infections was only 61, number of deaths was 9 and the number of recovered was 30. To overcome the difficulty in visualizing the statistics of first four weeks, we divided the data into two folds-first five weeks and second five weeks. Separate coxcomb diagrams were then constructed for each group.

Figure 4 shows the coxcomb diagram for the data of first five weeks. This graph is clearer and easier to understand as compared to the previous graph (i.e. Figure 3). This coxcomb graph is like a pie chart, cut into five equal angles each representing a week. These slices advance in a clockwise direction. Each shows what happened in one week of the COVID-19 period. The outward reach of each slice shows how many infections per crore, deaths per crore and recoveries per crore occurred in that week represented by the areas shaded in red, green and blue, respectively. It is important to keep in mind that the area is measured from the centre and the shaded areas are overlaid and not stacked. We see small slices in the first, second, third and fourth weeks since the rates were small during the earlier period of the outbreak. However the fifth slice, representing the fifth week, reached far outward in the radial direction-meaning much more infections were detected in the fifth week (maximum about 6 per crore) as compared to the first four weeks. For the second and third slices representing the third and fourth weeks, the blue shaded areas were much larger than the green shaded areas indicating that the recovery rates were large in these weeks relative to the death rates. But the situation reversed in the fifth week, which is when the death rate was much higher than the recovery rate — an indication that the outbreak was taking a deadly turn.

**Figure 3:**
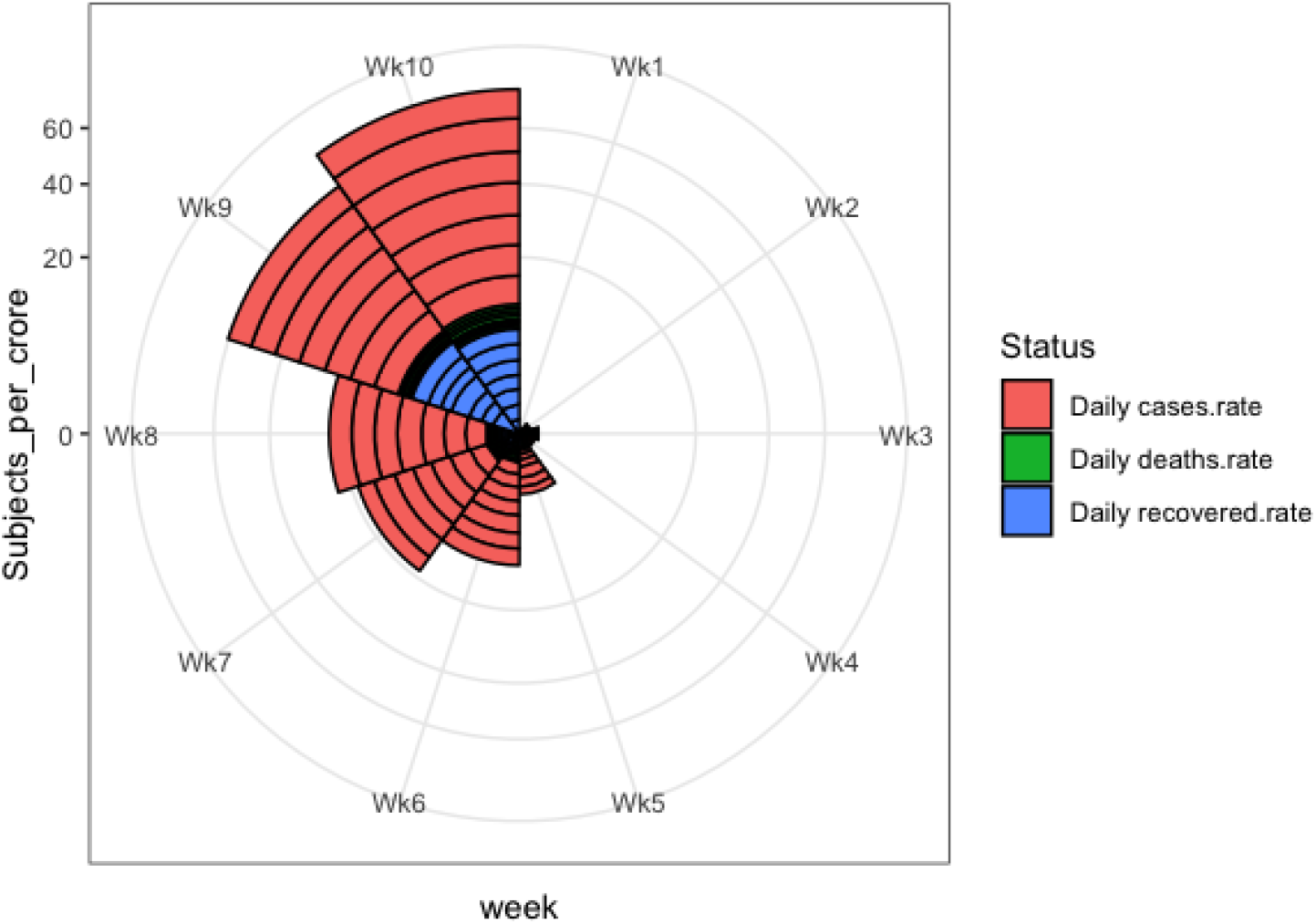
Coxcomb diagram for daily rate of infections, deaths and recovery for Bangladesh for 10 weeks since March 8, 2020.

Figure 5 shows the coxcomb diagram for the data of the second five-week period (April 12-May 16). This graph is even easier to understand than the previous graph (i.e. Figure 4). In this coxcomb graph, the outermost reach of each slice shows how many infections per crore occurred in that week. The radius and henceforth the area of each slice increases as we progress in the clockwise direction indicating increase in the rates of infection in successive weeks. The slices representing the 6th, 7th and 8th weeks are noticeably smaller in size than that of the 9th and 10th weeks. The coxcomb shows that there was a big jump in the infection rate from week 8 to week 9. The largest slice was for the 10th week meaning much more infections were detected during this time (i.e. maximum infection rate about 70 per crore) followed by the ninth week (i.e. maximum infection rate nearly 50 per crore). The death rates and recovery rates also appear to be considerably higher in the 9th and 10th weeks as compared to weeks 6, 7 and 8.

**Figure 4:**
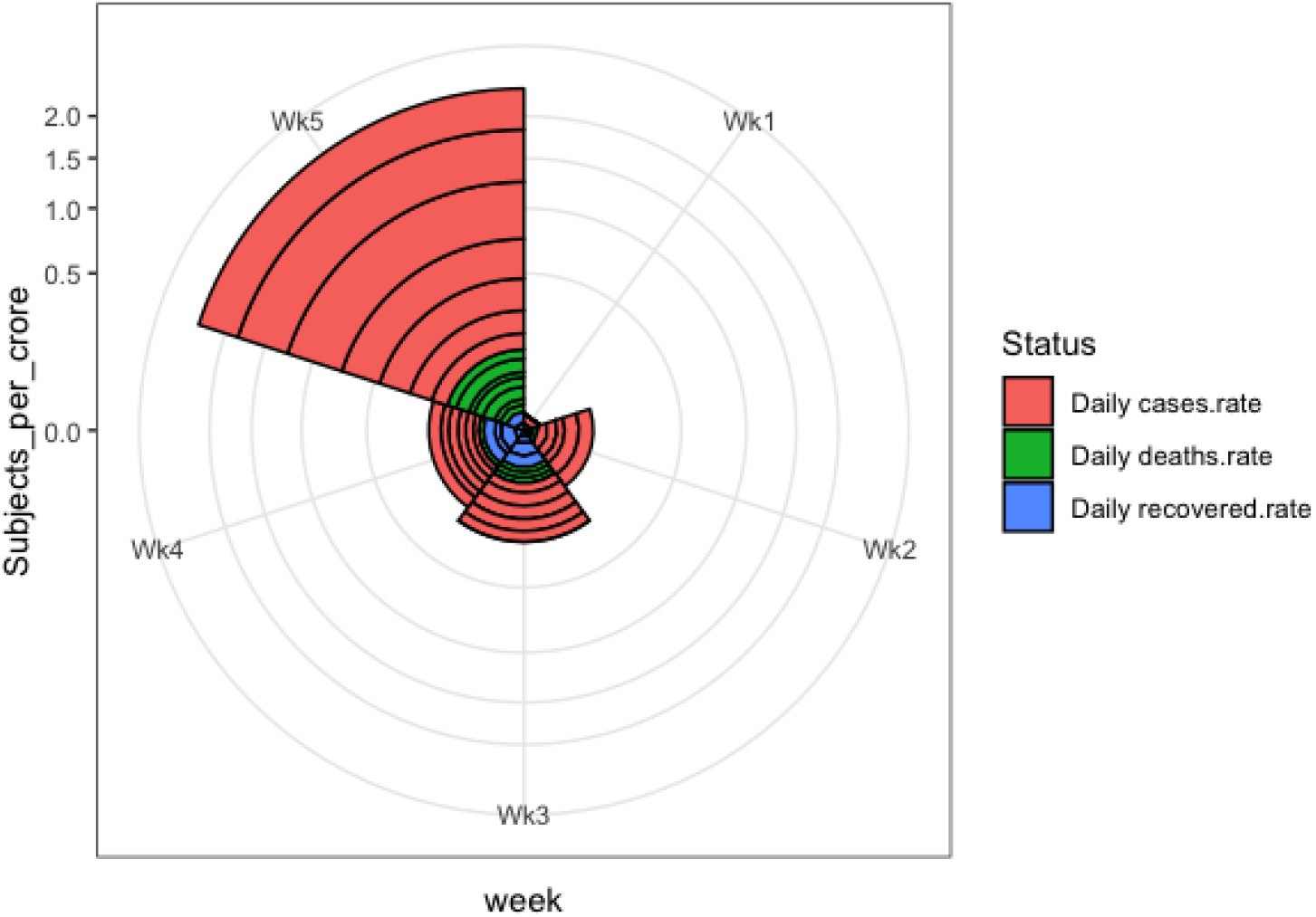
Coxcomb diagram for daily rate of infections, deaths and recovery for Bangladesh for week 1 to 5.

**Figure 5:**
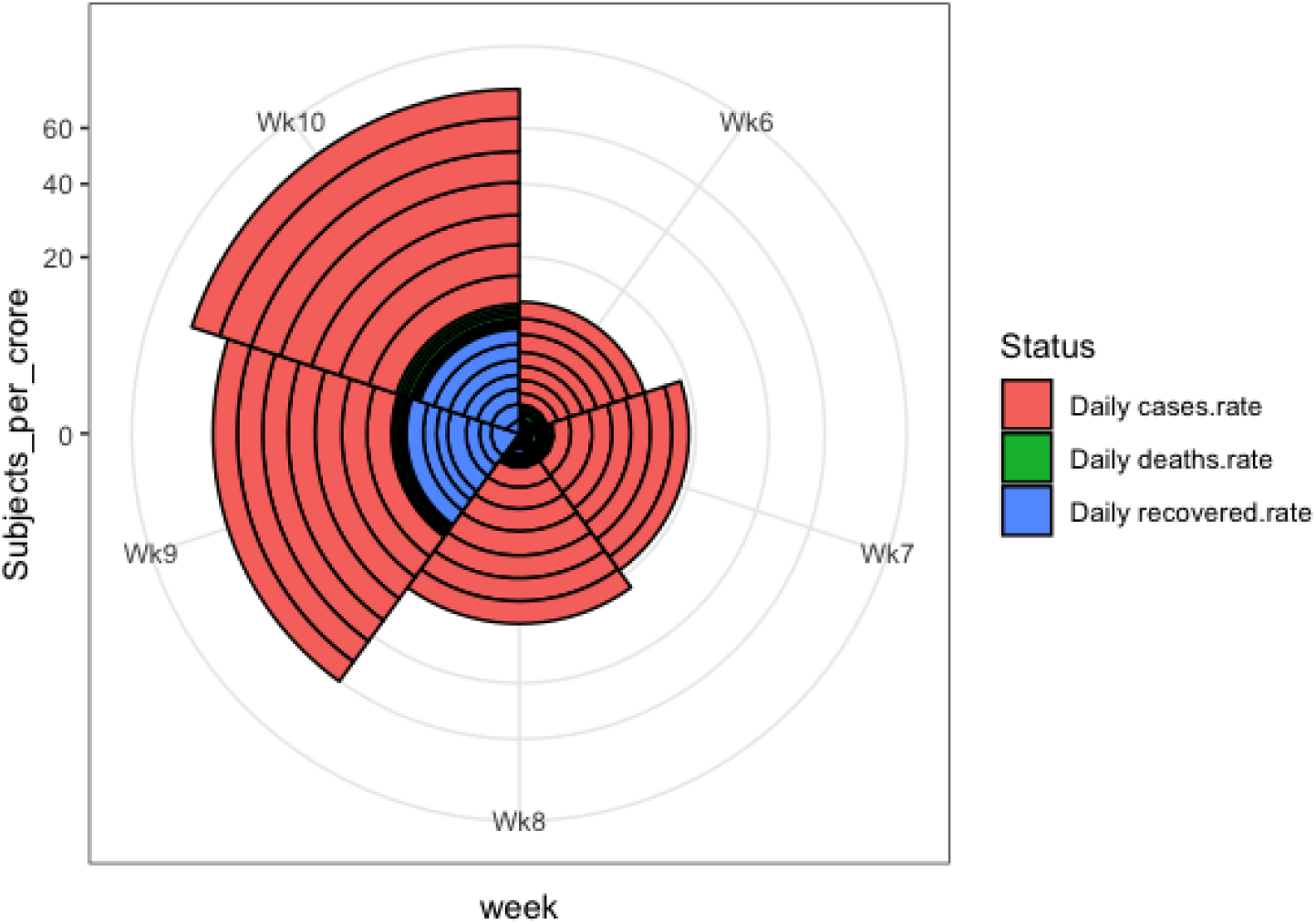
Coxcomb diagram for daily rate of infections, deaths and recovery for Bangladesh for week 6 to 10.

As it appears from these graphs Nightingale could, for example, have scaled deaths according to the radius, instead of the area, of the segments. That would’ve strengthened her case. But it would have misled people, since area is what the eye sees. *Hence we could use a Florence Nightingale today, as we drown in more undifferentiated data than anyone could have imagined during the Crimean War*— *could not understand needs to be reworded*. Similar to the above pictorial displays, coxcomb graphs could be generated for other COVID-19 rates such as the daily active rate, daily test rate and daily case finding rate of tests. In addtion, separate coxcomb graphs could be generated for all the rates according to various socio-demogrraphic differentials like urban-rural, male-female etc. In the next section, we demonstrate how the coxcomb graphs are generated by using the statistical language `R`.

### 3.2 Coxcomb Graphs Using R

The data used for this analysis is called BDcorona.xlsx, which is an excel dataset. However, any other format can be used depending on the user’s interests. This dataset has been extracted from the worldometer website (Max Roser & Ortiz-Ospina, 2020). This dataset is also available with this manuscript as a supplementary file. We assume the user is familiar with R studio platform. First, the following packages need to be installed along with the ‘readxl’ package for reading excel dataset. The dataset must then be loaded from the respective drive.

`library(knitr)`

`library(tidyverse)`

`library(magrittr)`

`library(grid)`

`library(gridExtra)`

`library(readxl)`

`bdcor <- read_excel(“~/BDcorona.xlsx”,col_names = TRUE,col_types = NULL)`

The following code may then be used to select the subset of data for which the coxcomb graphs in Figures 4 and 5 will be created. In our case, we will create coxcombs using the variables ‘Daily deaths.rate’, which represents the rate of daily reported deaths per crore, ‘Daily recovered.rate’, which represents the rate of daily recovered individuals per crore, and ‘Daily cases.rate’, which represents the rate of daily reported infections per crore. We present here the first 5 rows of the selected subset named as ‘coronabd’.

`kable(bdcor)`

`coronabd <- bdcor %>%`

`as_tibble %>%`

`subset(select = c(1, 7:9)`

`kable (coronabd)`

| \|Date | \| Daily deaths.rate\| | Daily recovered.rate\| | Daily cases.rate\| |
| --- | --- | --- | --- |
| :--- | ---: | ---: | ---: |
| \|2020-03-08 \| | 0.0000000\| | 0.0000000\| | 0.1764706\| |
| \|2020-03-09 \| | 0.0000000\| | 0.0000000\| | 0.0000000\| |
| \|2020-03-10 \| | 0.0000000\| | 0.0000000\| | 0.0000000\| |
| \|2020-03-11 \| | 0.0000000\| | 0.0000000\| | 0.0000000\| |
| \|2020-03-12 \| | 0.0000000\| | 0.1176471\| | 0.0000000\| |

Next, we describe the code that will be used to update the dataset ‘coronabd’ by adding the week and group information. Recall that our data contains information up to week 10. We wish to split the weeks into two groups called ‘Before’, which represents weeks 1 to 5 (from March 8 to April 11) and ‘After’, which represents weeks 6 to 10 (April 12 to May 16). The following code will lead to the desired data format. The first and last 3 rows of the updated dataset are shown below.

`coronabd %<>%`

`gather(key = “Status”, value = “Subjects_per_crore”, -Date) %>%`

`mutate(week = gl(10, 7, 210, labels = c(“Wk1”, “Wk2”, “Wk3”,”Wk4”, “Wk5”,”Wk6”,”Wk7”,”Wk8”,”Wk9”,”Wk10”))) %>%`

`mutate(Regime = gl(2, 35, 210, labels = c(“Before”, “After”), ordered = TRUE))`

`kable(coronabd)`

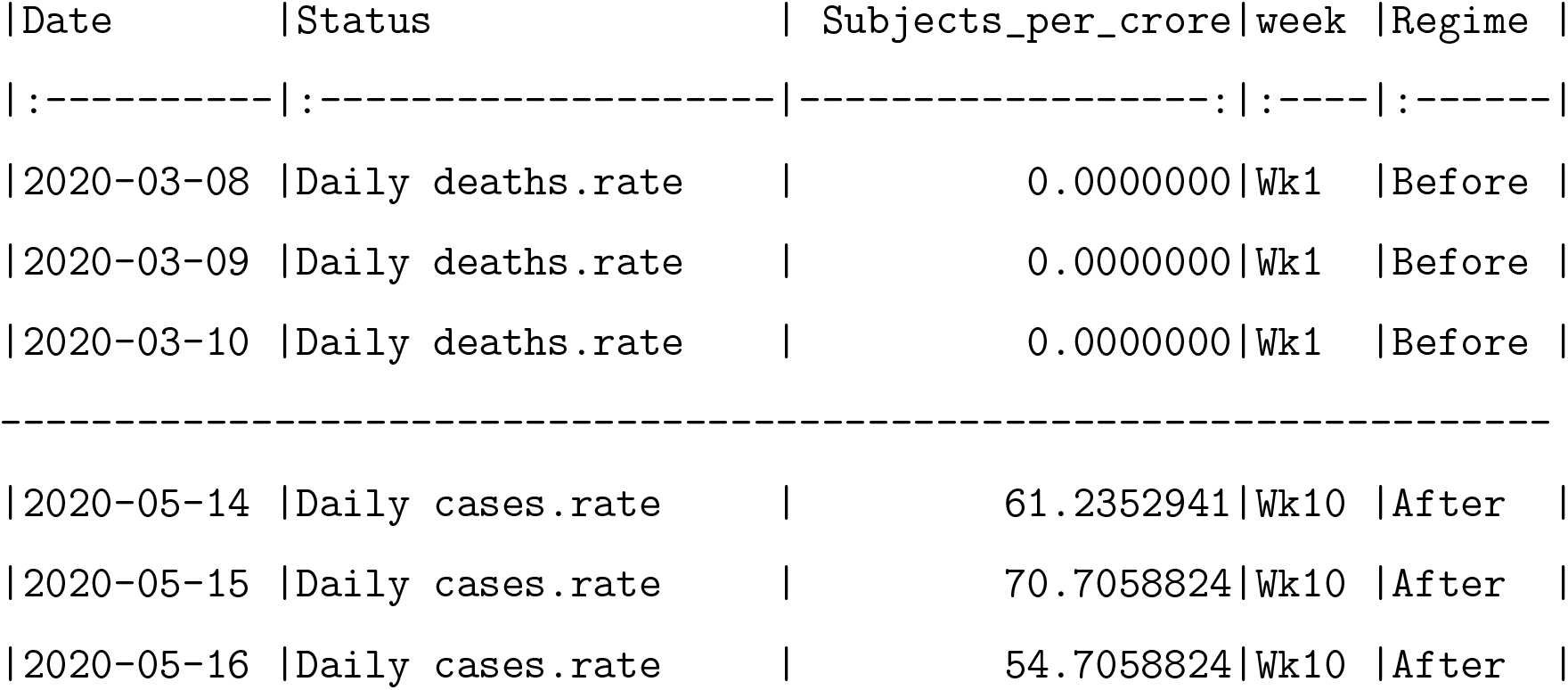

Then, we update the above ‘coronabd’ dataset with the type of variables by using following code. For our case, we have four non-numeric variables and one numeric variable. We also label here the ‘Status’ variable with the main three variables (‘Daily deaths’, ‘Daily recovered’, ‘Daily cases’) that lead to three rate variables.

`str(coronabd)`

`coronabd$Status %<>%`

`coronabd$Status %<>%`

`factor(levels = c(“Daily deaths”, “Daily recovered”, “Daily cases”))`

`coronabd %>%`

`kable(align = c(“c”, “c”, “r”, “c”, “c”))`

The following code produces the stacked barplot, Figure 2, as shown in this manuscript.

`cxc_b1 <- ggplot(data = coronabd,`

`aes(x = factor(Date), y = Subjects_per_crore, fill = Status)) +`

`geom_bar(width = 0.8, stat = “identity”, position = “stack”, colour = “black”) +`

`theme(axis.text.x = element_text(angle = 90, hjust = 1),`

`axis.ticks.x = element_blank(),`

`legend.position = c(0.3, 0.8)) +`

`scale_fill_brewer(type = “qual”, palette = “Pastel1”)`

`cxc_b1`

The following code produces the coxcomb graph, Figure 4, as shown in this manuscript for the group ‘Before’ that represents weeks 1 to 5.

`cxc11 <- ggplot(data = coronabd %>% subset(Regime == “Before”),`

`aes(x = week, y = Subjects_per_crore, fill = Status)) +`

`geom_bar(width = 1, stat = “identity”, position = “stack”, colour = “black”) +`

`scale_y_sqrt() +`

`coord_polar()`

`cxc11`

Similarly, the following code produces the coxcomb graph, Figure 5, as shown in this manuscript for the group ‘After’ that represents weeks 6 to 10.

`cxc22 <- ggplot(data = coronabd %>% subset(Regime == “After”),`

`aes(x = week, y = Subjects_per_crore, fill = Status)) +`

`geom_bar(width = 1, stat = “identity”, position = “stack”, colour = “black”) +`

`scale_y_sqrt() +`

`coord_polar()`

`cxc22`

The graphs in Figures 4 and 5 generated by the above code should show the rate per crore. Yet it is seen that the y-axis shows the rates expressed as a percentage. If one does not wish to see the rate expressed as a percentage on the y-axis, then the rates in the raw dataset need to be divided by 100. We do not show here the codes for generating the combined graph i..e. Figure 2 since it may be obtained through a slight modification of the codes given above.

## 4 Conclusions

This article, which aims at understanding the trend of the COVID-19 pandemic in Bangladadh, is a perfect example of how a 150 year old chart may be resurrected to understand the evolution of a 20th century pandemic that has rattled the world. The Florence Nightingale diagram, popularly known as the coxcomb, has been used in this article to display COVID-19 data in Bangladesh. The graph has been constructed for three major variables, namely, the rates of infections, deaths and recovered persons all expressed in per crore of the population. We found the coxcomb graph to be useful in visualizing the relative changes of the variables when the segments were considered to be weeks. Since the detection of the first COVID-19 case in early March, the epidemic appeared to be under control with very low rates of infection over the next four weeks. However, the graph clearly showed a rise in the rate of infections after the fourth week and rapidly increasing numbers of infections in successive weeks up till the 10th week.

A jump in the rate of infections from the previous week was apparent for weeks 5 and 9, which seems to indicate that social distancing measures had yet not made a dent in the spread of the virus even in later stages of the outbreak. By splitting the ten-week period into two five-week periods and creating separate coxcombs for each, it was possible to obseve the relative changes in the variables over time with greater detail. For instance, it was easy to discern that the rate of daily deaths per crore surpassed the rate of daily recoveries per crore in week 5. Thus, the coxcomb enabled one to compare the relative change in the numbers in a particular week relative to other weeks easily. However, if the pandemic persists for a long period, one might benefit from using the segments to represent months since the relative changes would be more apparent in this case. In contrast to the stacked barplot, which is generally constructed for daily data, the coxcomb appears to be more appealing because the former can sometimes be difficult to understand when the time series is long and the daily counts are very small. This article has also described the codes to reproduce the coxcomb graphs shown in the figures using popular statistical language R. By providing the codes, it is hoped that researchers will feel inspired to construct coxcomb diagrams of their own and in doing so pay tribute to a legend of statistics and data visualization and an architect of modern nursing.

In a nice write-up in the Guardian (Hoyos, Accessed May 15, 2020), the writer imagined what Nightingale would do if she were alive today battling the COVID-19 war. She wrote,‘Were Nightingale alive today, she would not be walking, torch in hand, among the patients of the Covid-19 hospitals named after her. She would instead be gazing intently at her laptop, her smartphone holding thousands of texts with the most influential people of the day, from the Queen and the prime minister to mathematicians and epidemiologists. Her computer would be filled with data-laden spreadsheets and she would be having a lively Twitter debate about the reliability of death figures.’ Nightingale is not amongst us today, but her legacy lives on as we learn to tell the story in the data using data visualization techniques and pursue her philosophy of making data-driven decisions.

## Data Availability

The working data set used for this study has been submitted to the journal as additional supporting file.

